## Supplemental Figures and Tables for "Single-cell analysis of psoriasis resolution reveals an inflammatory fibroblast state targeted by IL-23 blockade"

**Supplementary Figures**

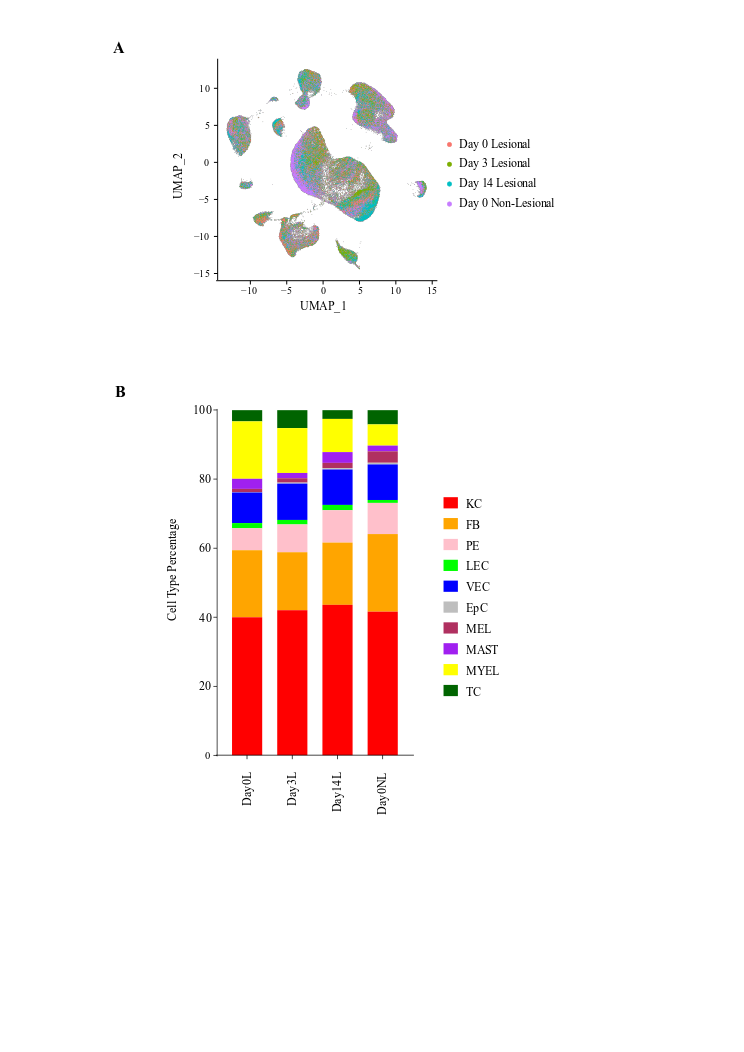

**Supplementary Figure 1: Clustering and abundance of cell populations detected in treated and untreated skin**. (**A**) Unifold Manifold Approximation and Projection (UMAP) of 164,741 single cells showing that cluster membership is not influenced by sample type (lesional vs non-lesional skin) or study time point. (**B**) Stacked bar chart showing the abundance of the various cell populations in the four sample groups. L, lesional skin; NL, non-lesional skin.

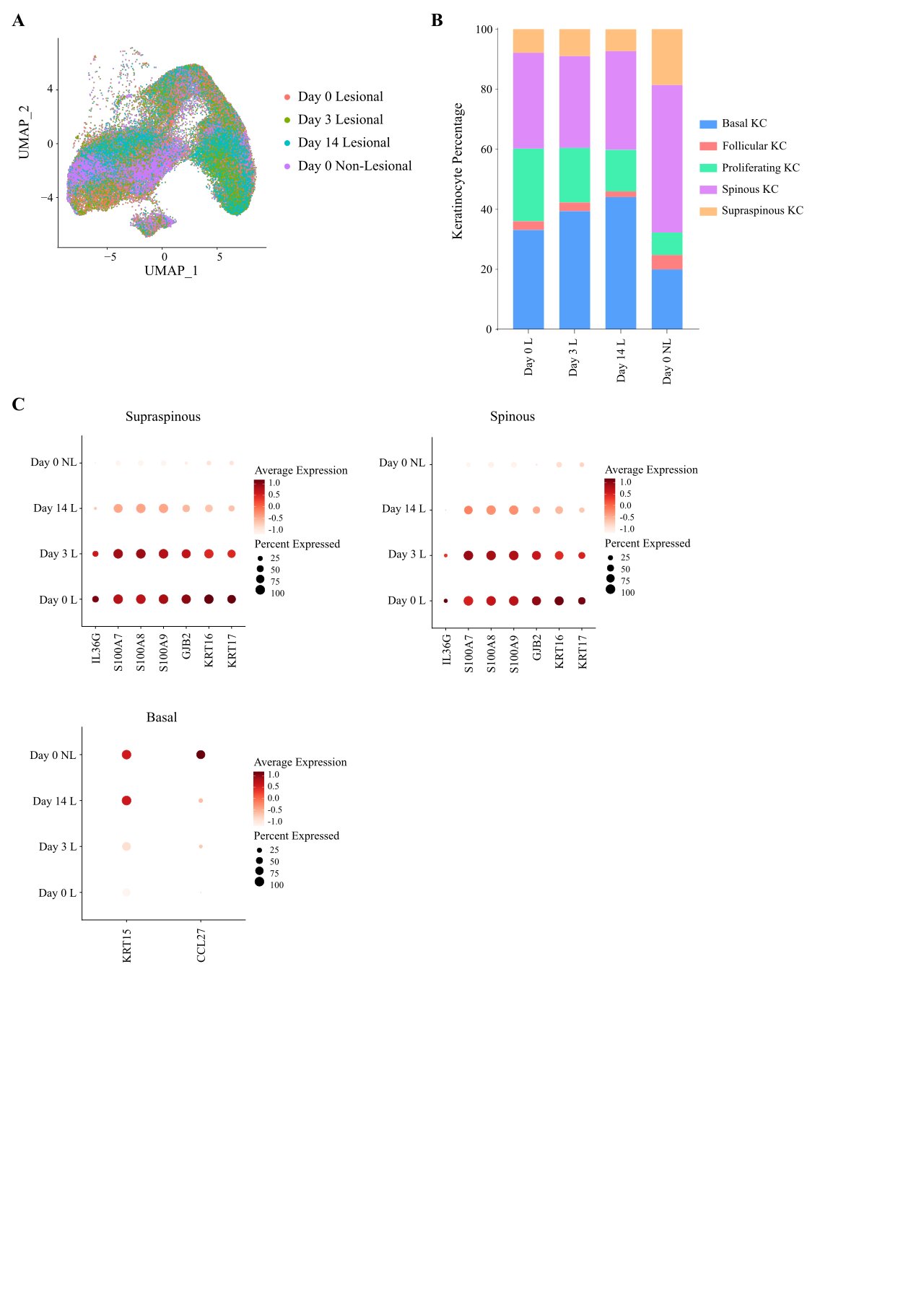

**Supplementary Figure 2: Clustering and abundance of keratinocyte populations detected in treated and untreated skin.** (**A**) UMAP of 68,695 keratinocytes showing that cluster membership is not influenced by sample type (lesional vs non-lesional skin) or study time point. (**B**) Stacked bar chart showing the abundance of the various keratinocyte populations in the four sample groups. (**C**) Dot plots showing treatment-induced gene expression changes within keratinocyte populations. KC, keratinocytes; L, lesional skin; NL, non-lesional skin.

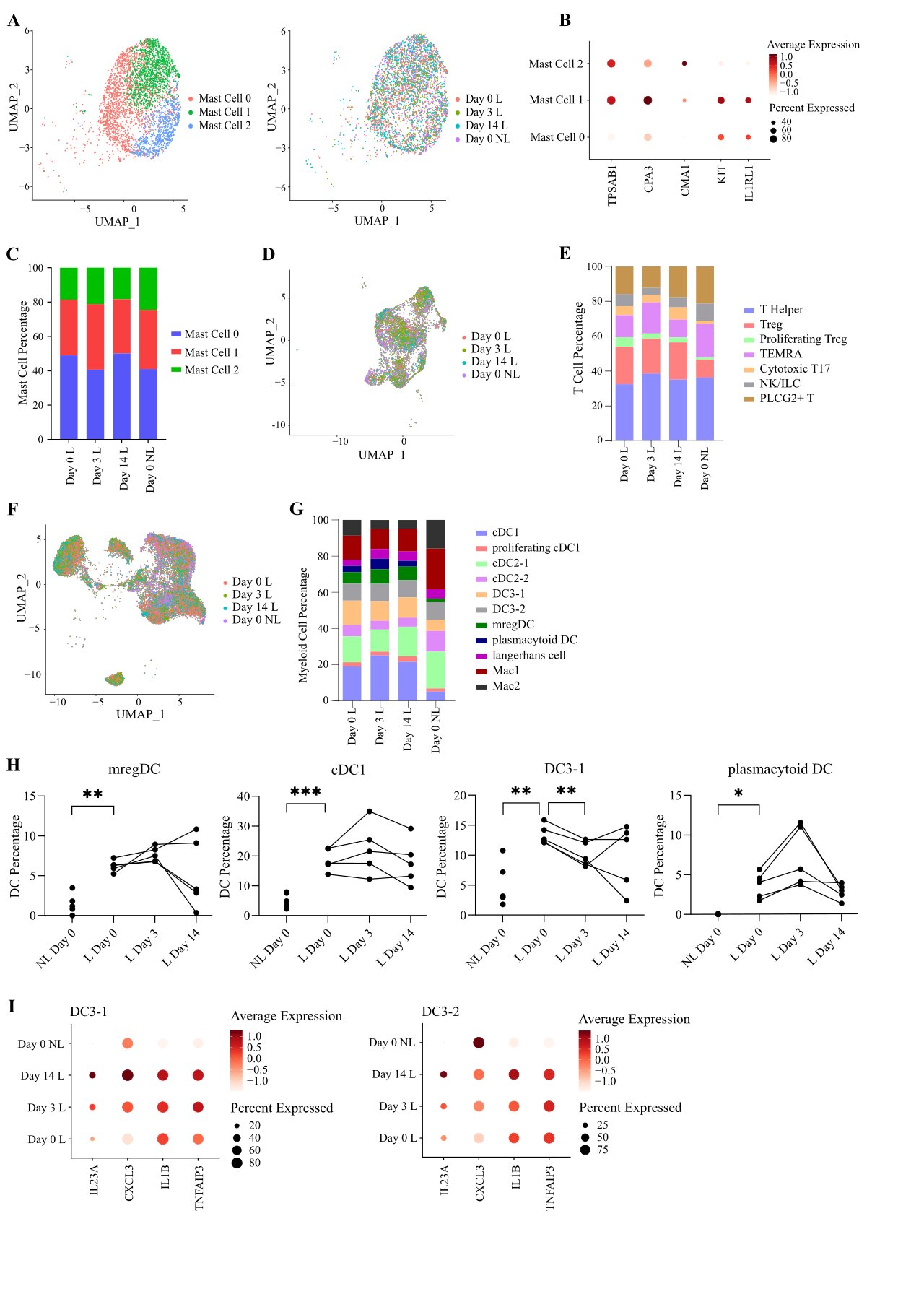

**Supplementary Figure 3: Clustering and abundance of immune populations detected in treated and untreated skin.** (**A**) UMAP of 3,755 mast cells forming three distinct sub-clusters. Cells are coloured by sub-cluster (left) or sample group (right). (**B**) Dot plot showing the expression of marker genes in the three sub-clusters. (**C**) Stacked bar chart showing the abundance of the various mast cell populations in the four sample groups. (**D**) UMAP of 6,263 T cells showing that cluster membership is not influenced by sample type (lesional vs non-lesional skin) or study time point. (**E**) Stacked bar chart showing the abundance of the various T cell populations in the four sample groups. (**F**) UMAP of 18,544 myeloid cells showing that cluster membership is not influenced by sample type (lesional vs non-lesional skin) or study time point. (**G**) Stacked bar chart showing the abundance of the various myeloid populations in the four sample groups. (**H**) Plot showing the abundance of myeloid populations at different time points. Every line represents a patient. **P*<0.05, **P*<0.01, **P*<0.001 (repeated measures ANOVA with Dunnett’s post-test). (**I**) Dot plots showing treatment-induced gene expression changes within DC3-1 (left) and DC3-2 (right) cells. L, lesional skin; NL, non-lesional skin; TEMRA, terminally differentiated effector memory T cells.

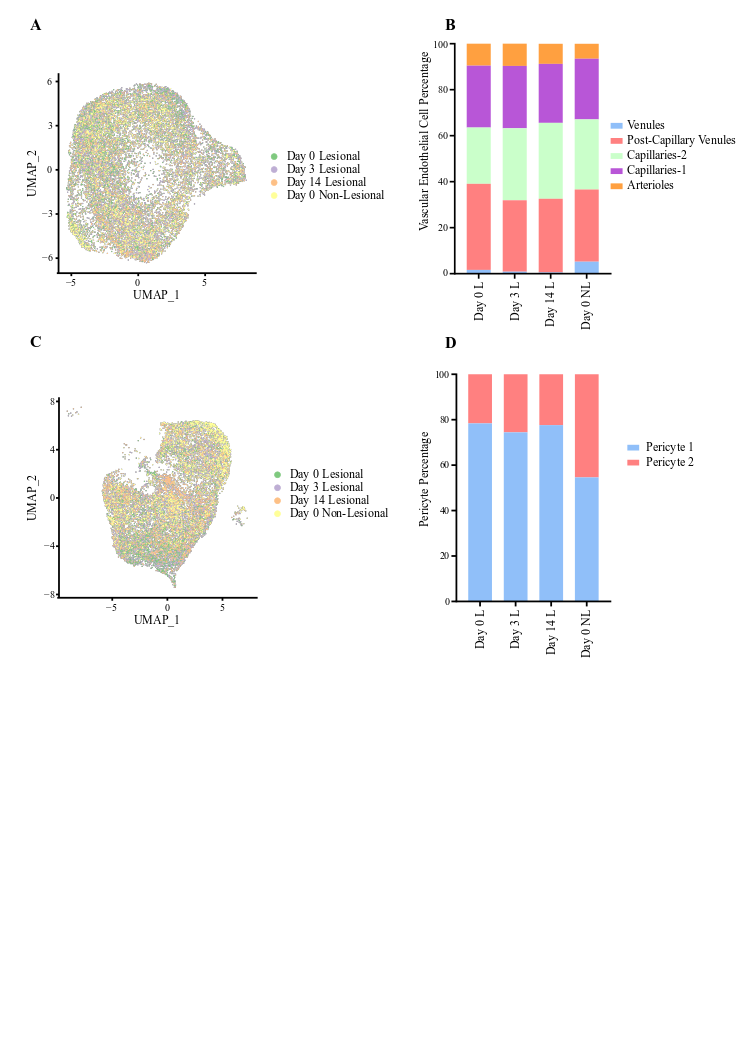

**Supplementary Figure 4**: **Clustering and abundance of vascular endothelial cells and pericytes in treated and untreated skin**. (**A**) UMAP of 16,420 vascular endothelial cells showing that cluster membership is not influenced by sample type (lesional vs non-lesional skin) or study time point. (**B**) Stacked bar chart showing the abundance of the various vascular endothelial cell populations in the four sample groups. (**C**) UMAP of 13,507 pericytes showing that cluster membership is not influenced by sample type (lesional vs non-lesional skin) or study time point. (**D**) Stacked bar chart showing the abundance of the various pericyte populations in the four sample groups.

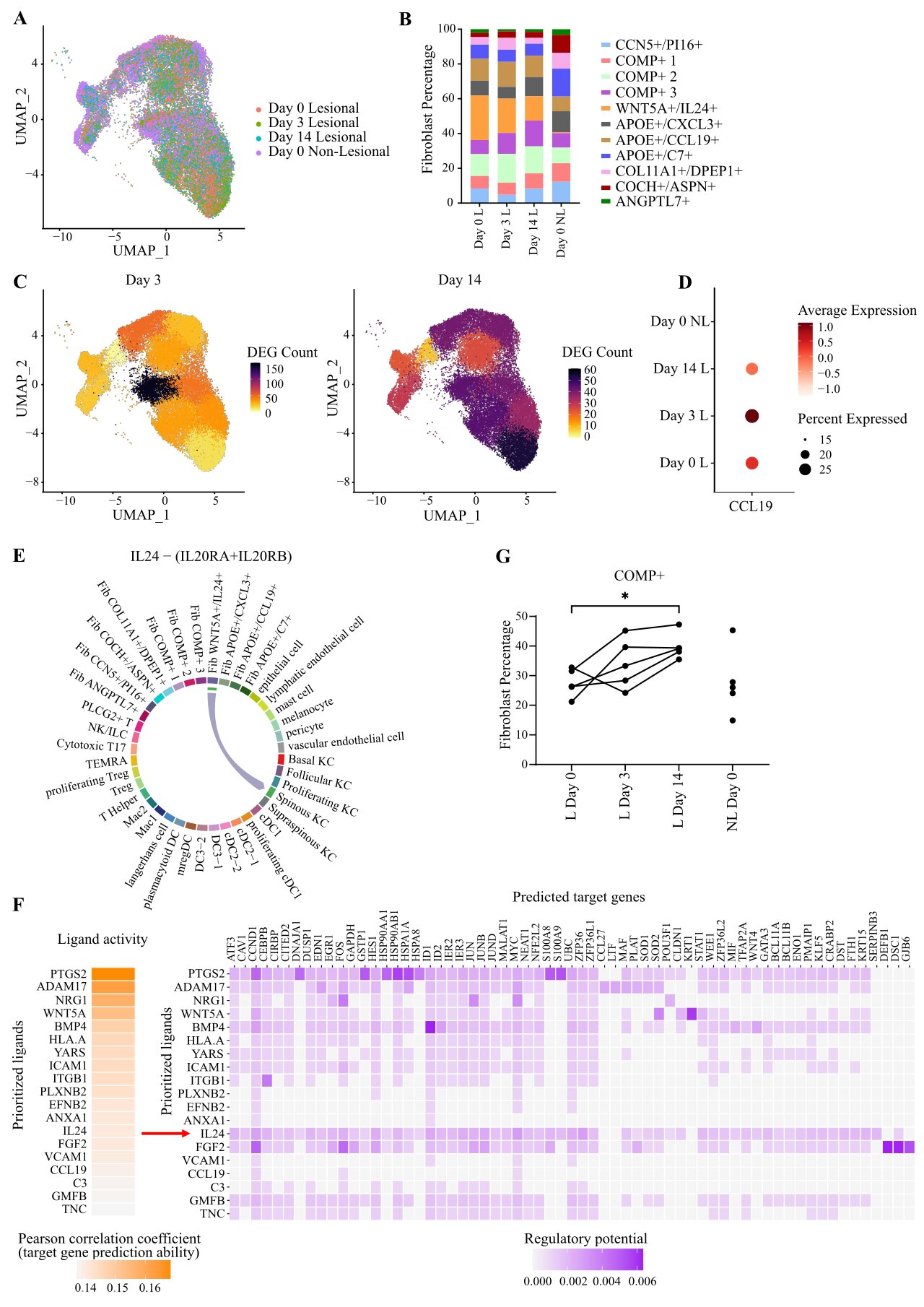

**Supplementary Figure 5**: **Clustering and abundance of fibroblast populations detected in treated and untreated skin.** (**A**) UMAP of 31,765 fibroblasts showing that cluster membership is not influenced by sample type (lesional vs non-lesional skin) or study time point. (**B**) Stacked bar chart showing the abundance of the various fibroblast populations in the four sample groups. (**C**) UMAP visualization of fibroblasts from lesional skin, showing the number of DEG observed in each cluster, after 3 (left) and 14 (right) days of treatment. (**D**) Dot plot showing the expression of *CCL19* in fibroblasts, following treatment with risankizumab. (**E**) Inferred interaction between IL-24 produced by *WNT5A*+/*IL24*+ fibroblasts and its receptor (encoded by *IL20RA* and *IL20RB*) on spinous keratinocytes. (**F**) NicheNet ligand activity prediction for the communication between *WNT5A+/IL24+* fibroblasts and spinous keratinocytes. Left: potential ligands expressed by *WNT5A+/IL24+* fibroblasts ranked according to their ability (Pearson correlation coefficient) to predict the gene expression changes observed between day 0 and day 14. Right: matrix showing the potential targets of the top ranked ligands and their regulatory potential in spinous keratinocytes. (**G**) Plot showing the abundance of COMP+ fibroblasts at different time points. Every line represents a patient. Fib, fibroblasts; L, lesional skin; NL, non-lesional skin; **P*<0.05 (repeated measures ANOVA with Dunnett’s post-test).

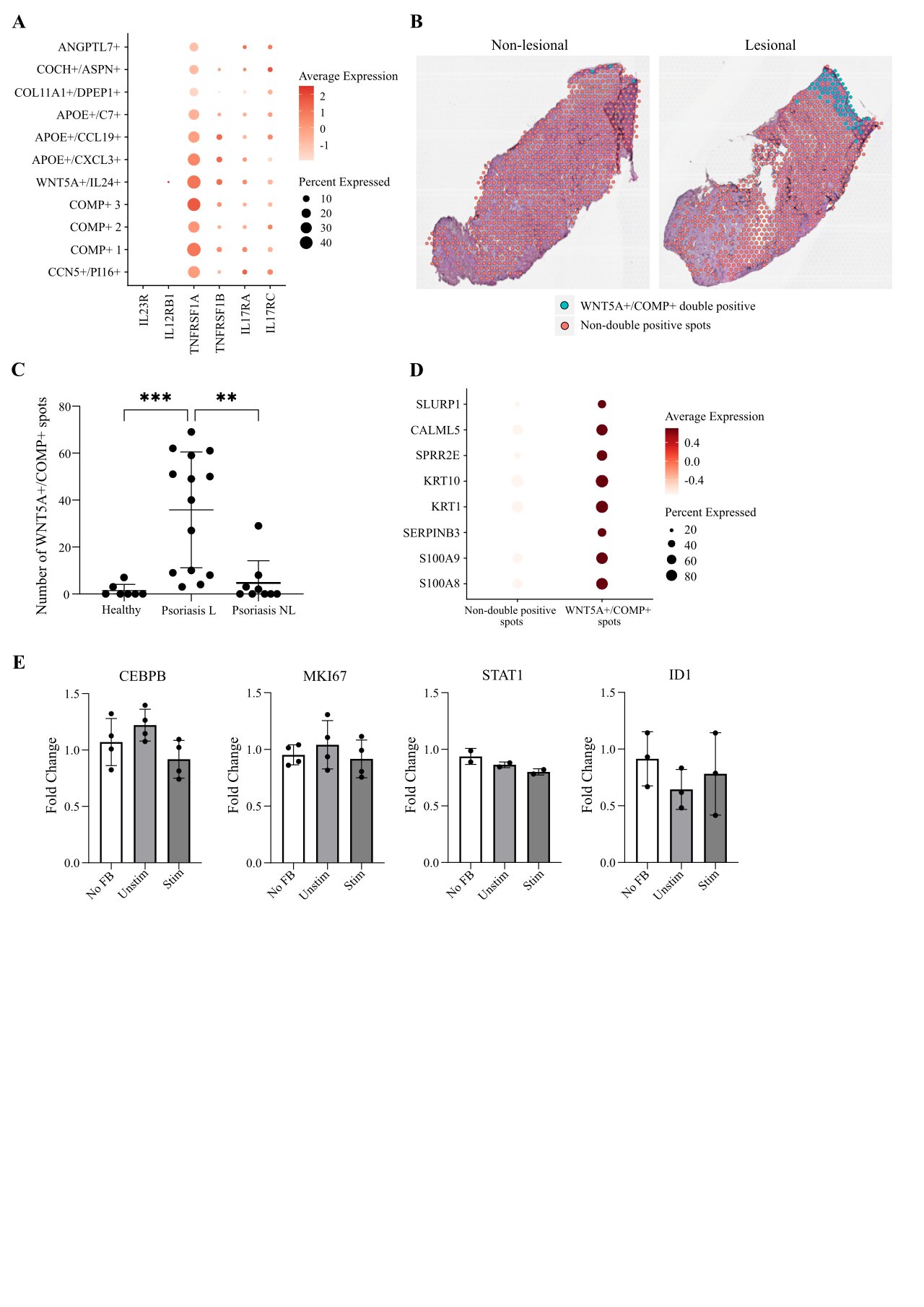

**Supplementary Figure 6**: **Spatial localization of *WNT5A+/IL24+* fibroblasts and interacting partners.** (**A**) Dot plot showing the expression of immune receptor genes in the various fibroblast populations. (**B**) Representative spatial plots from non-lesional and lesional psoriasis skin showing the localization of *WNT5A+/COMP+* spots. (**C**) Number of *WNT5A+/COMP+* spots across sample groups. L, lesional skin; NL, non-lesional skin; ***P*<0.01; ****P*<0.001 (ANOVA with Dunnett’s post-test). (**D**) Dot plot showing the expression of spinous/supra-spinous keratinocyte markers in *WNT5A+/COMP+* spots vs all other spots. (**E**) Real time PCR analysis of inflammatory markers in human primary keratinocytes cultured with unconditioned medium (no FB grown), supernatants from IL-17A/TNF stimulated fibroblasts (Stim) or control medium (supernatant from unstimulated fibroblasts, Unstim), N=2-4.

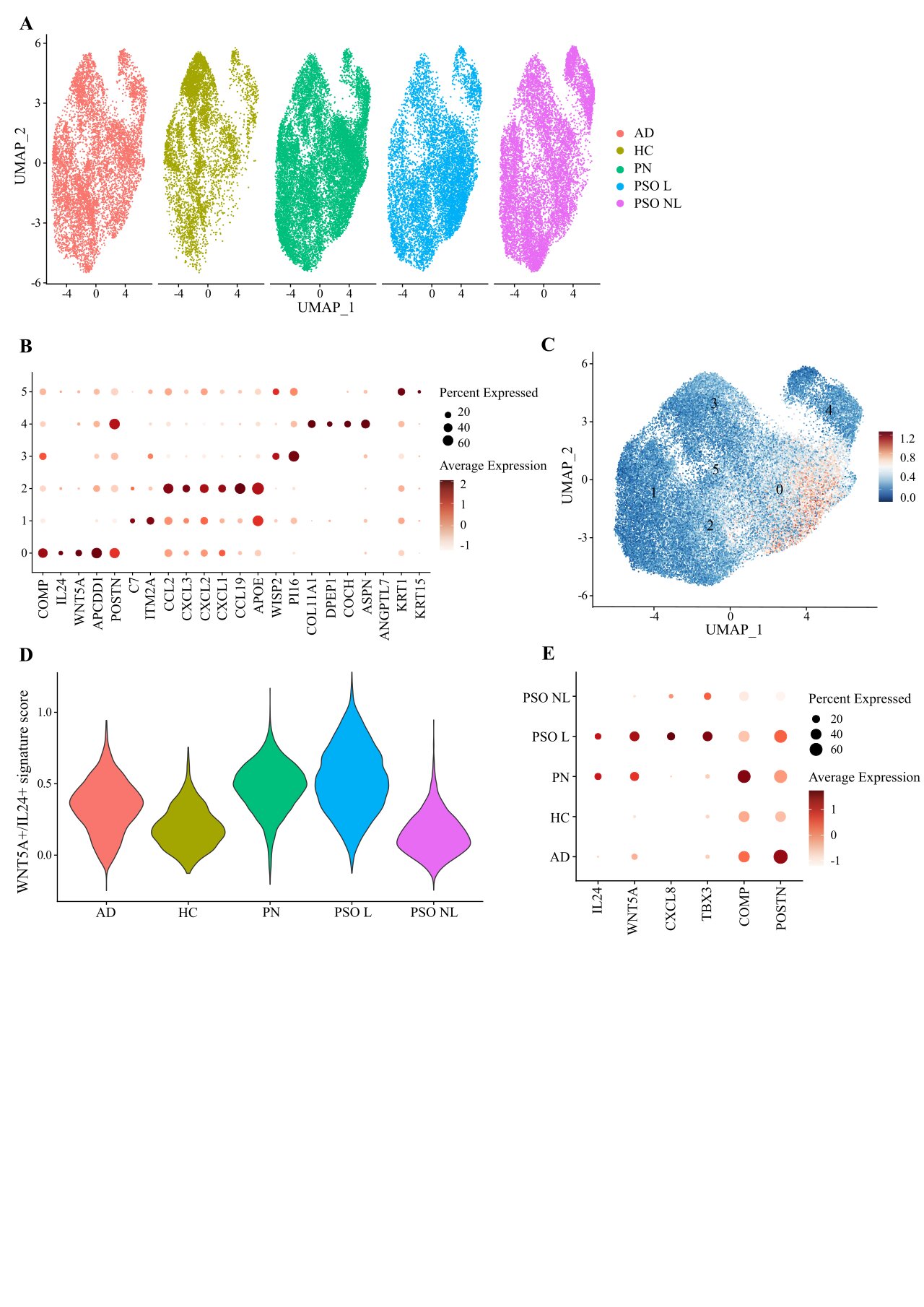

**Supplementary Figure 7**: **Analysis of** ***WNT5A+/IL24+* fibroblasts across inflammatory skin diseases.** (**A**) UMAP of fibroblasts from publicly available scRNA-seq datasets of healthy, prurigo nodularis and atopic dermatitis skin, following integration with our day 0 psoriasis data. Cells are coloured by sample group. (**B**) Dot plot showing the expression of marker genes used for the annotation of fibroblast sub-clusters. The *WNT5A+/IL24+* population corresponds to cluster 0 (**C**) UMAP of fibroblasts populations confirming that the *WNT5A+/IL24+* cell signature is specific to cluster 0. (**D**) Violin plot showing the *WNT5A+/IL24+* signature score across sample groups. (**E**) Dot plot showing gene expression in *WNT5A+/IL24+* fibroblasts across sample groups. PSO, psoriasis; L, lesional skin; NL, non-lesional skin; HC, healthy control; PN, prurigo nodularis; atopic dermatitis, AD.

**Supplementary Tables**

**Supplementary Table 1: Patient demographics^1^**

|  | *Patient n.* | *Mean age*  *(SD)* | *Sex* | *Comorbidity* | *Mean baseline PASI (SD)* | *Biologic naïve* |
| --- | --- | --- | --- | --- | --- | --- |
| Discovery  (scRNA-seq) | 5 | 30.1  (9.6) | 5M | None | 23.8  (9.9) | Yes |
| Validation  (RNA-scope) | 3 | 41.3  (10.1) | 2M  1F | None | 27.7  (11.7) | Yes |

^1^All individuals were of European descent; M, male; F, female; PASI, psoriasis area and severity index.

**Supplementary Table 2: scRNA-seq output summary statistics**

| *Sample Group* | *Mean n. of cells per sample (SD)* | *Mean n. of reads per cell (SD)* | *Mean n. of genes per cell (SD)* |
| --- | --- | --- | --- |
| Non-lesional skin  Day 0 | 8970 (1325) | 6967 (4427) | 1900 (776) |
| Lesional skin  Day 0 | 8382 (1583) | 7134 (5384) | 1953 (921) |
| Lesional skin  Day 3 | 7906 (1522) | 7138 (5552) | 1886 (902) |
| Lesional skin  Day 14 | 7653 (2384) | 7022 (4779) | 1944 (875) |

**Supplementary Table 3: Real-time PCR primers**

| *Target* | *Primer sequences (5’ to 3’)* |
| --- | --- |
| *CEBPB* | AACTCTCTGCTTCTCCCTCTG  AAGCCCGTAGGAACATCTTT |
| *CXCL8* | GAGAAGTTTTTGAAGAGGGCTGA  CTTCACTGATTCTTGGATACCACA |
| *GAPDH^1^* | CGGAGTCAACGGATTTGGTC  AATGAAGGGGTCATTGATGGCA |
| *ID1* | CAGTTGGAGCTGAACTCGGA  AACGCATGCCGCCTCG |
| *IL24* | TGTGGACTTTAGCCAGACCC  GGTAAAACCCAGGCAAGGGA |
| *MKI67* | CTTTGGGTGCGACTTGACGA  ACAACTCTTCCACTGGGACG |
| *PKG1^1^* | GCGGGTCGTTATGAGAGTCG  TGGGACAGCAGCCTTAATCC |
| *S100A8* | TTTCAGAAGACCTGGTGGGG  CAGGGAGTACTTGTGGTAGACG |
| *S100A9* | TCCTCGGCTTTGACAGAGTG  TGGTCTCTATGTTGCGTTCCA |
| *SERPINB3* | ACCAATGTGGTATTGCTGCCAA  AACTCCTGGGTGGAAAGTCAA |
| *STAT1* | TCTGTGTCTGAAGTTCACCCT  TCCGAGACACCTCGTCAAAC |
| *TBX3* | TGCACCTGGAGGCTAAAGAAC  GGAAACATTCGCCTTCCCGA |
| *WNT5A* | AGGGCTCCTACGAGAGTGCT  GACACCCCATGGCACTTG |

^1^Housekeeping genes
